# Biallelic WDR91 variants cause a neurodevelopmental disorder through impaired endosomal maturation and autophagy dysregulation

**DOI:** 10.64898/2026.04.03.26349989

**Authors:** Nathalie Merillon, Magalie Barth, Alban Ziegler, Patrick Van Bogaert, Sophie Gueden, Estelle Colin, Agnès Guichet, Gwendal Poussereau, Mélissa Giroudoux, Franck Geneviève, Isabelle Pellier, Coralie Mallebranche, Yves Delneste, Céline Beauvillain, Charline Miot

## Abstract

Biallelic variants in genes regulating endosomal, lysosomal and autophagy pathways are increasingly implicated in severe neurodevelopmental disorders, yet the contribution of the Rab7 effector WDR91 to human disease remains incompletely defined. We report a child with a severe neurodevelopmental disorder characterized by progressive microcephaly, microlissencephaly, corpus callosum hypoplasia, and early-onset epilepsy, harboring compound heterozygous *WDR91* variants: a truncating variant (p.Gln215*) and a missense variant (p.Tyr15Asn).

Functional analyses show that p.Gln215* abolishes WDR91 expression, whereas p.Tyr15Asn reduces protein abundance through increased degradation. Reduced WDR91 expression was confirmed in primary patient-derived cells. *In silico* analyses suggest that p.Tyr15Asn induces a localized change within an N-terminal degron-containing region, potentially affecting protein stability.

In cellular models, both variants impair early-to-late endosomal maturation and alter WDR91 localization to Rab7-positive compartments. WDR91 deficiency is further associated with transcriptional and functional evidence of autophagy dysregulation. While the p.Tyr15Asn variant partially restores autophagic flux under overexpression conditions, patient-derived cells display impaired autophagic turnover, consistent with a context-dependent functional and partial loss of function effect of this variant.

Together, these findings provide functional evidence supporting the pathogenicity of *WDR91* variants and implicate combined defects in endosomal maturation and autophagy in WDR91-related neurodevelopmental disease.

## Introduction

Malformations of cortical development, including congenital microcephaly and lissencephaly, represent major causes of severe early-onset neurodevelopmental disorders. These entities form a biological continuum reflecting disruption of core processes of corticogenesis, such as neural progenitor proliferation, neuronal migration, and cortical organization^1^. Postnatal brain growth may also be affected by impaired neuronal maintenance, membrane trafficking, and lysosomal function, highlighting the contribution of intracellular homeostasis pathways to neurodevelopmental disease^2^.

*WDR91* encodes a WD repeat-containing Rab7 effector that regulates endosomal maturation and lysosomal homeostasis. It is recruited to endosomes by active Rab7 and, in complex with WDR81, controls PI3K activity required for early-to-late endosome conversion^3^. WDR91 is highly expressed in the brain and is essential for neuronal endosomal trafficking and autophagy–lysosome function^4; 5^. In mice, loss of Wdr91 leads to impaired postnatal brain growth, defective endosomal maturation, altered dendritic development, and neuronal loss, supporting its critical role in neurodevelopment^4; 5^.

Recent reports have described children with severe neurodevelopmental phenotypes associated with biallelic *WDR91* variants, suggesting a recurrent clinical association^6^;^7^. However, these observations remained descriptive and lacked functional validation, leaving the causal role of WDR91 unresolved.

Here, we report a child with a severe neurodevelopmental disorder carrying biallelic *WDR91* variants and provide functional evidence supporting pathogenicity. Using cellular models and patient-derived primary cells, we show that WDR91 deficiency impairs endosomal maturation and disrupts autophagic homeostasis. These data establish a mechanistic link between WDR91 dysfunction and human neurodevelopmental disease.

## Subjects, material and methods

### Patient and controls

The proband and both parents were enrolled in the ORIGIN study (Functional Study to Identify the Genetic Etiology of Rare Diseases; ClinicalTrials.gov identifier NCT05499091; ID RCB 2022-A00473-40). The study protocol was approved by the appropriate institutional review board and conducted in accordance with the Declaration of Helsinki and relevant national regulations. Written informed consent was obtained from the parents prior to inclusion. The ORIGIN study establishes a biocollection to support functional validation of NGS-identified variants and to investigate their relationship with clinical phenotypes. Peripheral blood samples were collected from the proband and both parents for functional analyses.

Peripheral blood samples from healthy control donors were obtained through the Blood Collection Center (EFS Pays de la Loire, France; agreement ANG-2017-01) following written informed consent and approval by the local ethics committee (agreement CPDL-PLER-2021 038).

### Exome sequencing

DNA of patient and his parents was extracted from peripheral blood cells. Exome capture was performed using the Twist Human Core Exome Enrichment System (Twist Bioscience), and sequencing was performed on an Illumina HiSeq4000 platform. Base calling was performed using the Real-Time Analysis software sequence pipeline (2.7.6) with default parameters. Sequence reads were mapped to the human genome build (GRCh38) using Elandv2e (Illumina; CASAVA1.8.2) allowing multiseed and gapped alignments. Variant filtering was performed to consider variants affecting either coding sequences or splice site regions. High-quality variants were filtered against public (dbSNP150 and gnomAD V.2.0.1, MAF threshold ≤ 0.01% or unknown frequency for dominant and X-linked transmission and <0.1% for recessive transmission). After stratification taking into account genotype, inheritance model, and predicted functional impact, the different heterozygous variants of *WDR91* were considered as the best candidate genes underlying the trait. Validation and segregation of variants in *WDR91* were then assessed by Sanger sequencing.

### In silico analyses

Orthologous WDR91 protein sequences were retrieved from UniProt for human (*Homo sapiens*), mouse (*Mus musculus*), rat (*Rattus norvegicus*), zebrafish (*Danio rerio*), and *Caenorhabditis elegans*, with additional vertebrate orthologs included when available. Multiple sequence alignments were generated using Clustal Omega (v2.10)^8^ with default parameters and visualized in Jalview (v2.11.5.1)^9–11^, where residue-level conservation scores were computed using standard conservation metrics and similarity-based coloring schemes. Sequence logos for the N-terminal region (residues 1-60) were generated using WebLogo to illustrate conservation at the p.Tyr15 position. Nucleotide-level evolutionary constraint at the codon underlying p.Tyr15 was assessed using PhyloP and PhastCons scores from the UCSC Genome Browser (100-way vertebrate alignments, GRCh38), focusing on *WDR91* exon 1.

The three-dimensional structure of wild-type WDR91 (UniProt A4D1P6-1) was obtained from the AlphaFold Protein Structure Database. The p.Tyr15Asn substitution was introduced in silico using PyMOL (v3.1.6.1; Schrödinger, LLC), followed by energy minimization and stability analysis using FoldX (v5.1). Structural superposition, root-mean-square deviation (RMSD), and ΔΔG stability calculations were performed in PyMOL and FoldX.

Post-translational modification (PTM) predictions were performed on the full-length sequence using NetPhos 3.1 (phosphorylation)^12; 13^, ASEB (lysine acetylation)^14^, and GPS-UbER 5.0 (ubiquitination)^15^, with independent validation by MusiteDeep^16–18^. Predicted sites were mapped onto the multiple sequence alignment in Jalview to assess evolutionary conservation.

Intrinsic disorder and disorder-mediated binding propensity were predicted using IUPred2A (long mode) and ANCHOR2 under identical parameters for WT and p.Tyr15Asn WDR91 sequences^19; 20^. Residue-level scores (0 to 1) were compared based on absolute differences and local averages surrounding the variant, as these algorithms do not provide statistical outputs. The N-terminal region was additionally screened for degron motifs using DEGRONOPEDIA^21; 22^, and predicted degrons were interpreted in the context of local disorder and ANCHOR profiles.

### Generation of WDR91 Knockout Cell Lines

WDR91 knockout (KO) HEK293T cell lines were generated using CRISPR/Cas9-mediated genome editing. Guide RNAs (gRNAs) targeting exons 1 (51-CGCCGAGATCAAGGCGGACA-31) and 5 (51-TGTCTCCAACATGGACCGCC-31) of the human *WDR91* gene were cloned into the pX330-U6-Chimeric_BB-CBh-hSpCas9 vector (gift from Feng Zhang; Addgene plasmid #42230), as previously described^23^; ^24^. HEK293T cells were co-transfected with the two CRISPR constructs together with a reporter plasmid encoding human CD4 (pcDNA3.1-Hygro-delta-hCD4; gift from Craig Bassing, Children’s Hospital of Philadelphia, University of Pennsylvania, PA) using Lipofectamine 2000 (Thermo Fisher Scientific) according to the manufacturer’s instructions. Twenty-four hours post-transfection, CD4⁺ cells were isolated by cell sorting (FACSAria II; BD Biosciences) and single-cell cloned. Clonal populations were expanded for approximately two weeks and screened for *WDR91* gene inactivation through Western blotting.

### Lentiviral constructs and cells transduction

A lentiviral expression vector encoding the human *WDR91* open reading frame (NM_001362737.2) under a CMV promoter and a tagBFP2 reporter gene under a EF1A promoter (VectorBuilder) was used as the wild-type construct. Site-directed mutagenesis was performed using the QuikChange II XL Site-Directed Mutagenesis Kit (Agilent Technologies) to generate the indicated mutant variants. All constructs were sequence-verified prior to use. Lentiviral particles production was performed in a BSL-2 laboratory certified for vector production (University of Angers). Lentiviral particles were produced by transfecting HEK293T cells with the respective expression constructs together with packaging components using the Lenti-vpak Packaging Kit (Origene) according to the manufacturer’s instructions. Viral supernatants were collected at 72 hours post-transfection, filtered through 0.45 µm filters, and used immediately for transduction. For transduction, WDR91^KO^ HEK293T cells were subjected to spinoculation by centrifugation at 1,500×g for 90 min in 2 mL of viral supernatant supplemented with 8 µg/mL polybrene (Sigma-Aldrich). Cells were then incubated at 37°C for 5 days. Transduced tagBFP2-positive cells were isolated by cell sorting (FACSAria II) to generate stable, homogeneous cell populations. Control cell lines were generated using the corresponding empty lentiviral vector (VectorBuilder) and are referred to as WDR91^empty^ cells. WDR91 expression was validated by Western blotting and immunofluorescence. All HEK293 cell lines were cultured in DMEM medium supplemented with L-Glutamine, penicillin, streptomycin and fetal calf serum (10%). For some experiments, cells were treated with cycloheximide (10 μg/ml, Sigma) or MG132 (20 μMol, Sigma) for up to 4 hours.

### Western blotting

Cells were lysed in ice-cold lysis buffer (20 mM Tris-HCl, pH 7.5, 100 mM NaCl, 0.1% (w/v) SDS, 0.5% (w/v) sodium deoxycholate, 1 mM PMSF) supplemented with Complete Protease Inhibitor Cocktail (Roche). Protein concentrations were determined using standard colorimetric assays, and 20 µg of total protein per sample were resolved by SDS-PAGE on 4-12% gradient gels (ExpressPlus PAGE Gel; GenScript). Proteins were transferred onto PVDF membranes and incubated with the indicated primary antibodies (anti-WDR91 Ab, Abcam; all other primary antibodies, Cell Signaling Technology). Fluorescent detection was performed using IRDye 800CW Goat anti-Rabbit IgG and IRDye 680LT Goat anti-Mouse IgG2b Ab (LI-COR Biosciences). Membranes were imaged using an Odyssey infrared imaging system (LI-COR Biosciences). β-actin was used as the loading control. Signal quantification was performed using ImageJ (v1.52q, NIH). Multichannel images were split to isolate individual fluorescence channels corresponding to different secondary antibodies, accounting for channel-specific background differences. Each channel was converted to grayscale and inverted prior to measurement of band mean intensity. Protein levels were normalized to β-actin.

### Confocal microscopy - Immunofluorescence

HEK293T cells were cultured on glass coverslips in DMEM medium supplemented with FBS and penicillin/streptomycin, before being fixed using 4% (v/v) paraformaldehyde and permeabilized using PBS containing 0.1% (v/v) TritonX100. Cells were then saturated using TBS-Tween 0.01% supplemented with 3% goat serum, and stained with a primary anti-human WDR91 Ab (Abcam) and anti-human EEA1 or anti-human Rab7 Abs (Cell Signaling Technologies) and a secondary goat anti-mouse/rabbit IgG mAb coupled to AF488 or AF687 (Thermo Fisher Scientific). Nuclei were counterstained with SYTOX Orange (Invitrogen), and coverslips were mounted using ProLong Diamond Antifade Mountant (Molecular Probes). Images were acquired on a Leica DMI 6000 confocal fluorescence microscope and analyzed with ImageJ (v1.52q, NIH). For colocalization analyses, Z-stacks were collected at 0.2 μm intervals using a 63× objective with a zoom factor of 2.5 and 12-bit acquisition depth, resulting in a pixel size of 72 nm. Sequential scanning was performed to avoid spectral overlap. Laser power settings were adjusted to achieve comparable mean fluorescence intensities across channels. Colocalization was quantified using Pearson’s correlation coefficient to minimize bias related to fluorescence intensity differences.

### Endosomal maturation assay

HEK293T cells were cultured on glass slides in DMEM medium supplemented with 10% FBS and 1% penicillin/streptomycin prior to transfection with pmCherry-2xFYVE plasmid (gift from Harald Stenmark; Addgene plasmid #140050)^25; 26^ and GFP-rab7 WT plasmid (gift from Richard Pagano, Addgene plasmid #12605)^27^. Live-cell imaging was performed to monitor endosomal dynamics. Glass-bottom dishes were placed in a temperature-controlled, humidified chamber maintained at 37°C with 5% CO₂ (the Brick and The Cube 2 systems, Life Imaging Services). Time-lapse images were acquired every 30 sec for 60 min using a confocal fluorescence microscope (Leica DMI 6000). Z-stacks were collected at 0.4 µm intervals and acquisitions were performed with a 63x objective, a zoom factor of 3 for a pixel size of 60 nm. GFP and mCherry were excited at 488 nm and 561 nm, respectively. Image analysis was performed using ImageJ (v1.52q; NIH).

Early-to-late endosomal conversion time was defined at the single-vesicle level as the interval between the first detection of a 2×FYVE-mCherry-positive endosome and the acquisition of detectable GFP-Rab7 signal on the same vesicle, concomitant with loss or marked reduction of the FYVE signal. Vesicles were tracked manually across time points to ensure continuity. If Rab7 recruitment did not occur within the 60-min acquisition period, the conversion time was censored and recorded as 60 min. Conversion times from at least 10 vesicles per condition were pooled for statistical analysis.

### RNA-sequencing

Total RNA was purified using the RNeasy Plus microkit (Qiagen). mRNA libraries and sequencing were performed by IntegraGen and bioinformatics analyses were performed using Galileo software (IntegraGen). Briefly, mRNA libraries were prepared after polyA mRNA capture and Illumina indexes were added to cDNA using unique dual indexes. RNA-seq libraries were then subjected to paired-end 100 bp read sequencing on an Illumina NovaSeqX+ sequencer (70 million total reads per sample). Bioinformatic processing and analyses is detailed in the Supplementary material.

### Autophagy assay

HEK293T cells were cultured in DMEM medium supplemented with 10% FBS and 1% penicillin–streptomycin for 48 h prior to autophagy induction. Cells were then incubated in Hank’s Balanced Salt Solution (HBSS) for 2 h to induce nutrient deprivation, followed by treatment with 10 nM bafilomycin A_1_ Sigma Aldrich) for an additional 2 h to inhibit lysosomal degradation and enable assessment of autophagic flux. Subsequently, 2×10^5^ cells were stained with an autophagy detection probe (Autophagy Assay Kit; Bio-Rad) for 30 min at 37°C according to the manufacturer’s instructions. Cell viability was assessed using the LIVE/DEAD Fixable Near-IR Dead Cell Stain Kit (Invitrogen) prior to acquisition. For primary T lymphoblasts, the same protocol was applied except that cells were maintained in ImmunoCult medium (Stem Cell Technologies) supplemented with 10 ng/mL IL-2 without medium replacement prior to bafilomycin A_1_ treatment. Data were acquired on a CytoFLEX flow cytometer (Beckman Coulter) and analyzed using FlowJo software (v10.10.0; BD Biosciences). Dead cells were excluded prior to quantification. For each independent experiment, autophagy signal values were normalized to the lowest replicate (consistently WDR91^WT^ HEK293T cells or lowest healthy donor T cells) and expressed as fold change relative to WT/HD baseline to allow comparison across experiments and to account for inter-experimental variability.

### Primary T cells culture

Peripheral blood mononuclear cells (PBMCs) were isolated from whole blood by density gradient centrifugation using Ficoll-Paque. T cells were purified by magnetic sorting using the Human Pan T Cell Isolation Kit and LS columns (Miltenyi Biotec). T cell purity, assessed by flow cytometry, consistently exceeded 90%. Purified T cells were cultured in ImmunoCult medium supplemented with 5 ug/ml phytohemagglutinin (PHA-P; Sigma Aldrich) and 10 ng/mL recombinant human IL-2 (Stem Cell Technologies). Fresh rhIL-2 was added every 48 h to maintain T lymphoblasts survival. To sustain proliferation, T cells were restimulated every 8-10 days using 1 µg/mL anti-CD3 (clone OKT3) and 2 µg/mL anti-CD28 Ab (both from BioLegend). All functional assays were performed the day prior to scheduled TCR restimulation to assess baseline cellular function and minimize confounding effects related to acute TCR-induced signaling.

### Statistical analysis

All data was analyzed in GraphPad Prism 10 using statistical tests indicated in the figure legends. Error bars indicate mean +/- SEM unless otherwise stated. n.s., p>0.05; *p<0.05; **p<0.01; ***p<0.001; ****p<0.0001.

## Results

### Clinical findings

We report a boy who was first referred before one year of age for congenital microcephaly. At birth, he presented with severe microcephaly (−3 SD) with normal height and weight. Microcephaly progressed during infancy, reaching −7 SD after five years of age (Figure 1A). Severe epilepsy with multiple daily seizures developed within the first months of life. Brain MRI before one year of age revealed microlissencephaly with corpus callosum hypoplasia (Figure 1B). During follow-up, the patient developed treatment-resistant epilepsy and profound psychomotor impairment, with absence of head control, inability to sit, and no speech after five years of age. Serum biomarkers of neuronal and astrocytic injury showed increased neurofilament and GFAP levels (13.9 pg/mL; reference 5.20-10.8 and 30.8 pg/mL; reference 1.72-11, respectively).

**Figure 1:**
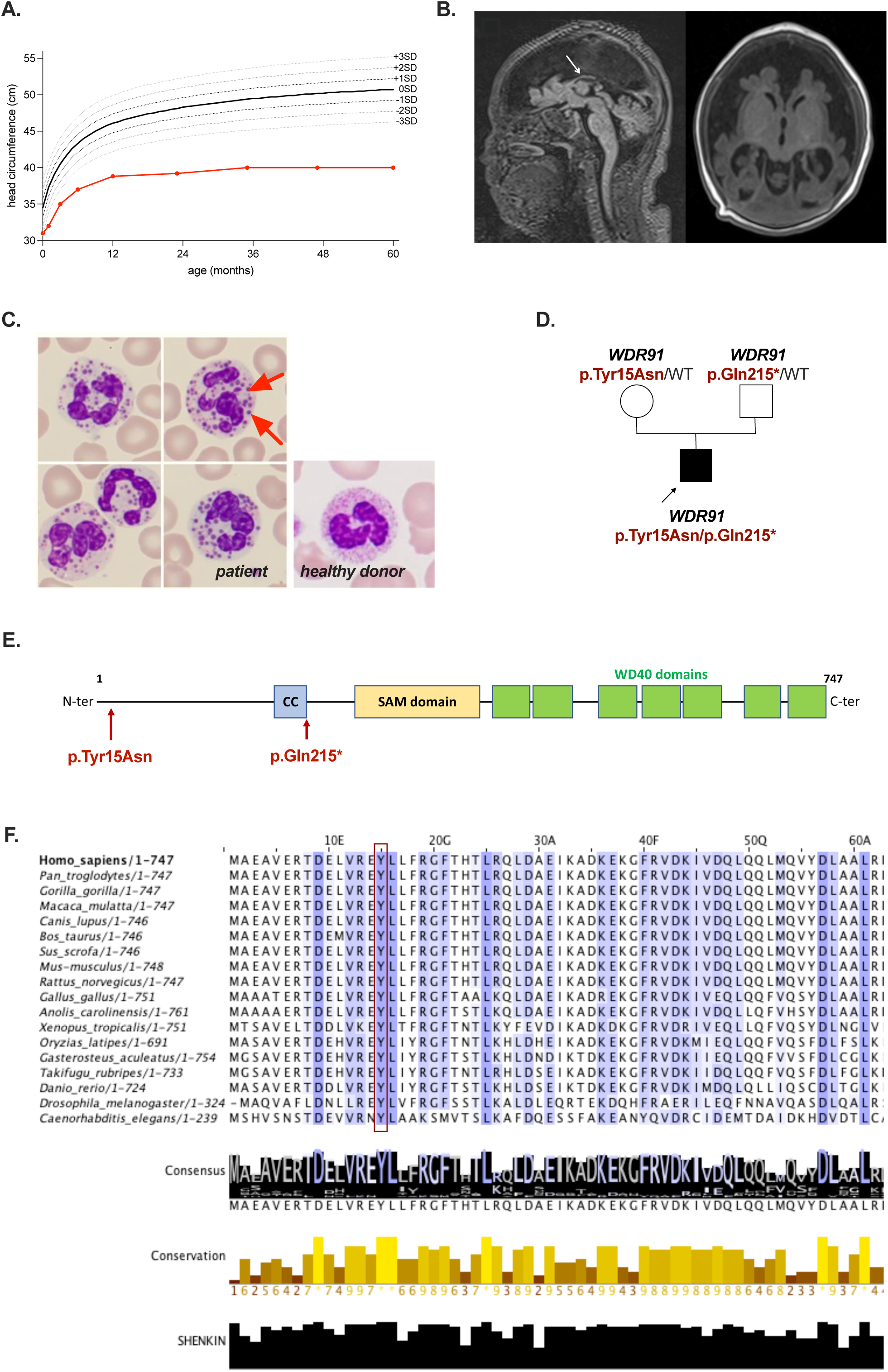
Clinical presentation and genetic findings. **A.** Diagram of head circumference since birth. **B.** T1 sequences of brain MRI before one year. On sagittal view (left), the corpus callosum is hypoplasic and incomplete (arrow). Brainstem and cerebellum appear normal. On axial view (right), the frontal and parietal cortex is profoundly hypoplasic with a simplified gyrus pattern. The operculum is widely open. These findings are consistent with microlissencephaly. **C.** Optical microscopy on peripheral blood smear after MGG staining, of patient and healthy donor. Images are centered on neutrophils. Giant granulations are prominent in the cytoplasm of patient’s cells (left panel, red arrows), whereas no abnormal granulation is visible in healthy donor’s cells (right). **D.** Pedigree of the family. WDR91 variants are indicated for each subject. **E.** WDR91 protein structure. Candidate variants are indicated along the protein (red arrows). **F.** Phylogenetics analysis of the N-terminal domain of WDR91 protein (amino acids 1-60).

Routine laboratory tests showed a normal blood count. However, blood smear analysis revealed giant cytoplasmic granules in neutrophils (Figure 1C). Such granules have been described in disorders affecting lysosome-related organelles, including Chediak-Higashi syndrome^28^. In the present case, the early and severe neurological presentation was not typical of this condition.

### Identification of compound heterozygous variants of WDR91 gene

Analysis of trio-based exome sequencing identified compound heterozygous variants in *WDR91* in the proband, comprising a maternally inherited missense variant (NM_001362736:c.43T>A; p.(Tyr15Asn)) and a paternally inherited predicted loss-of-function nonsense variant (c.643C>T; p.Gln215*) (Figure 1D). Both variants were absent from population reference databases, including the 1000 Genomes Project, dbSNP and gnomAD, supporting their rarity. No pathogenic or likely pathogenic variants were identified in genes associated with Chediak-Higashi syndrome (*LYST*) or Hermansky-Pudlak syndrome type 2 (*AP3B1*), excluding known genetic causes of giant neutrophil granules.

WDR91 encodes a 747-amino acid protein comprising an N-terminal region, a coiled-coil domain, a sterile alpha motif (SAM) domain, and a large C-terminal WD40-repeat domain essential for endosomal protein interactions (Figure 1E). The c.643C>T p.Gln215* nonsense variant is located in exon 5/15 and therefore likely to induce non-sense mediated decay (NMD). In case of NMD escape, the residual protein is predicted to lack the WD40-repeat domain, consistent with loss of function. The p.Tyr15Asn variant is predicted to be deleterious (CADD score 31; damaging predictions by REVEL, MutPred, SIFT, and PROVEAN; Table 1). Cross-species alignment demonstrated strict conservation of Tyr15 from vertebrates to invertebrates, with maximal conservation scores and no tolerated substitutions at this position (Figure 1G). Concordantly, the underlying nucleotide resides within a highly constrained genomic region, as indicated by elevated PhyloP and PhastCons scores in the 100-way vertebrate alignment (Supplemental Figure S1). Together, these data support strong evolutionary constraint at Tyr15 and the functional relevance of this residue.

**Table 1:** *In silico* prediction of pathogenicity for the identified *WDR91* variants. Predicted functional impact of the missense variant p.Tyr15Asn was assessed using multiple in silico tools, including CADD, REVEL, MutPred, SIFT, PROVEAN, and PolyPhen-2, all of which supported a deleterious effect. The truncating variant p.Gln215* was not evaluated by these tools, as they are primarily designed for missense variants. Scores are reported according to each tool’s recommended interpretation thresholds. In silico prediction of pathogenicity of the identified *WDR91* variants. *NA: non attributable*.

| WDR91 variant (NM_001362736) | c.43T>A | c.643C>T |
| --- | --- | --- |
| Protein variant | p.Tyr15Asn | p.Gln215* |
| Exon (of 15 exons) | 1 | 5 |
| CADD | 31 | NA |
| REVEL | Pathogenic (0.878) | NA |
| Mutpred | Pathogenic (0.663) | NA |
| SIFT | Pathogenic (0) | NA |
| PROVEAN | Pathogenic (-7.37) | NA |
| PolyPhen | Probably pathogenic (1) | NA |

### WDR91 p.Tyr15Asn variant reduces protein abundance through increased degradation

We first evaluated WDR91 protein expression in CRISPR/Cas9-generated WDR91 knocked-out HEK293T cells (hereafter referred to as WDR91^KO^) reconstituted with lentiviral expression vectors encoding WT, p.Gln215*, or p.Tyr15Asn WDR91 cDNA (hereafter referred to as WDR91^WT^, WDR91^Q215X^, and WDR91^Y15N^ cells, respectively) or an empty lentiviral vector as control (WDR91^empty^). Western blot analysis demonstrated complete absence of WDR91 protein in cells expressing p.Gln215*, whereas p.Tyr15Asn was detected at significantly reduced levels relative to WT (Figure 2A). Similar results were obtained following stable plasmid transfection on WDR91^KO^ cells (Supplemental Figure S3A), confirming that p.Gln215* abolishes detectable WDR91 protein expression and that p.Tyr15Asn reduces steady-state protein abundance. Importantly, primary CD3^+^ T lymphoblasts derived from the patient exhibited decreased endogenous WDR91 protein levels compared with healthy donor controls, consistent with the results obtained on mutant-expressing HEK293 cells (Figure 2B). Moreover, CHO cells transiently transfected with increasing doses of plasmids containing Y15N *WDR91* cDNA exhibited significantly lower WDR91 protein expression compared to cells transfected with WT plasmids (Supplemental S3C), suggesting that p.Tyr15Asn variant impacts WDR91 protein stability either through preventing WDR91 protein synthesis or through increasing WDR91 protein degradation or both. Consistently, comparable *WDR91* mRNA levels were observed in WDR91^WT^ and WDR91^Y15N^ cells, supporting impaired protein stability rather than altered transcription (Supplemental Figure S3B).

**Figure 2:**
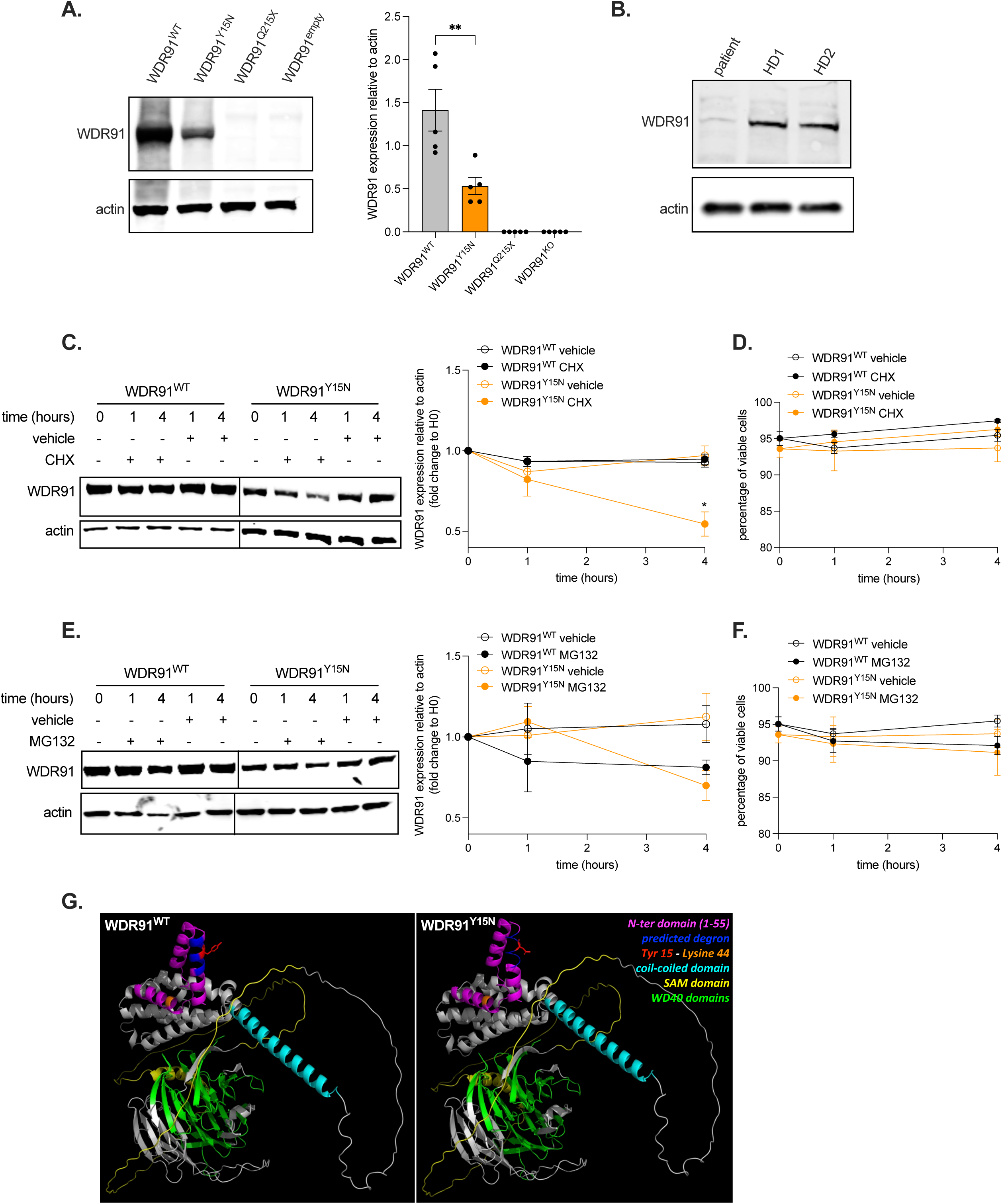
WDR91 p.Tyr15Asn variant reduces protein abundance through increased degradation. **A.** WDR91 protein expression in HEK293 WDR91^KO^ cells transduced with an empty lentiviral vector (WDR91^empty^) or a vector encoding WT (WDR91^WT^), p.Gln215* (WDR91^Q215X^), or p.Tyr15Asn (WDR91^Y15N^) WDR91 cDNA. Representative blot (left) and quantification of WDR91 expression relative to actin (right; Mann-Whitney test, * p<0.05). **B.** WDR91 protein expression in primary T cells from the patient and healthy donor (representative blot from 2 independent experiments). **C.** WDR91 protein expression in WDR91^WT^ and WDR91^Y15N^ cells after treatment with cycloheximide (CHX, 10 μg/ml) or vehicle (DMSO). Representative blot (left) and quantification of WDR91 expression relative to actin (right; 2-way ANOVA with Sidak multiple comparison test, * p<0.05). **D.** Viability of cells after cycloheximide treatment. **E.** WDR91 protein expression in WDR91^WT^ and WDR91^Y15N^ cells after treatment with MG132 (20 μM) or vehicle (DMSO). Representative blot (left) and quantification of WDR91 expression relative to actin (right). **F.** Viability of cells after MG132 treatment. **G.** AlphaFold-predicted WDR91 protein structure (left WDR91^WT^, right WDR91^Y15N^). Amino acid 15 residue is highlighted in red, known and predicted domains are highlighted in different colors. (**A,** C-F) All data collected are combined from three (**C-F**) to five (**A**) independent experiments. Bars indicate mean ± SEM. For Western blots, individual fluorescence channels were analyzed separately due to distinct background signal.

To determine whether reduced abundance resulted from altered synthesis or increased degradation, cells were treated with the proteasome inhibitor MG132 to inhibit protein degradation and thus assess protein synthesis, or with cycloheximide (CHX) to inhibit protein synthesis and thus assess protein degradation. Upon treatment with MG132, we did not observe any difference in WDR91 protein expression over time between WDR91^WT^ and WDR91^Y15N^ cells, indicating intact protein synthesis in WDR91^Y15N^ cells (Figure 2C). In contrast, CHX chase assays demonstrated accelerated decay of WDR91^Y15N^ compared to WT, with significantly lower protein levels after 4 h, confirming increased degradation kinetics (Figure 2E). Cell viability during MG132 and CHX treatment was comparable between genotypes, excluding differential cytotoxicity as a confounding factor (Figure 2D and 2F). Collectively, these results show that p.Gln215* results in complete loss of WDR91 protein, whereas p.Tyr15Asn reduces WDR91 abundance through increased protein degradation.

### In silico analyses of N-terminal determinants of WDR91^Y15N^ destabilization

To further investigate potential mechanisms underlying the increased degradation of WDR91^Y15N^, we analyzed predicted post-translational modifications within the N-terminal region. Although Tyr residues may serve as phosphorylation sites, NetPhos analysis did not support phosphorylation at Tyr15 (score < 0.5) (Supplemental Figure S4A). Similarly, ASEB-based prediction did not reveal a strong N-terminal acetylation signature, although a modest signal was observed for Lys36 under the GCN5/PCAF model (p < 0.1; Supplemental Figure S4B).

Prediction of lysine ubiquitination using GPS-UbER identified Lys33 and Lys44 as candidate sites, with identical outputs for WT and Y15N sequences. Independent analysis with MusiteDeep confirmed Lys44 (score 0.56) but not Lys33 as a candidate ubiquitination site, again with no differential ubiquitination predictions observed between genotypes. Given that degrons function as docking motifs for E3 ubiquitin ligases^29^, we examined potential degron sequences within the N-terminal region, specifically in the vicinity of Tyr15. Importantly, residues 11-18 encompass a sequence predicted to match an APC/C-associated D-box degron motif (Figure 2G). IUPred2A analysis revealed a modest increase in predicted intrinsic disorder across residues 1-65 in the p.Tyr15Asn variant relative to WT, as shown by elevated IUPred and ANCHOR scores (Supplemental Figure S4C). However, absolute intrinsic disorder scores remained below the conventional disorder threshold (IUPred > 0.5), indicating absence of a qualitative transition to a fully disordered state. Accordingly, structural modeling based on the AlphaFold-predicted WDR91 structure followed by FoldX energy calculations did not predict major global destabilization associated with p.Tyr15Asn, suggesting that the variant does not disrupt overall folding but may exert a localized functional effect within the highly conserved N-terminal region (Figure 2G and Supplemental Figure S2).

Together, these data suggest that the p.Tyr15Asn substitution induces a localized and modest change in the biophysical context of an N-terminal APC/C D-box degron motif, potentially modulating degron accessibility or recognition rather than causing major structural destabilization of the WDR91 N-terminus.

### WDR91^Q215X^ and WDR91^Y15N^ impair early-to-late endosomal maturation

As WDR91 loss has been described to impair early-to-late endosome conversion in HeLa cells, we examined endosomal maturation in the WDR91 mutant HEK293T cell lines using live-cell imaging. Endosomal conversion was monitored by tracking the disappearance of the early endosomal PtdIns3P reporter 2×FYVE-mCherry (2×FYVE-mCh) and the concomitant acquisition of the late endosomal marker GFP-Rab7.

In WDR91^WT^ cells, 2×FYVE-mCh signal rapidly disappeared while GFP-Rab7 was recruited to the same vesicles, consistent with efficient early-to-late endosomal conversion. In contrast, enlarged endosomal structures were observed in both WDR91^Q215X^ and WDR91^Y15N^ cells compared with WDR91^WT^ cells (Supplemental Figure 5), in line with previous observations in WDR91-deficient HeLa cells^4^. As expected, WDR91^Q215X^ cells exhibited a significant delay in conversion kinetics, consistent with loss of WDR91 function (Figure 3A). Despite detectable, albeit reduced, WDR91 protein levels in WDR91^Y15N^ cells, early-to-late endosomal conversion was also significantly delayed compared with WDR91^WT^ cells, with kinetics comparable to those observed in WDR91^Q215X^ cells (Figure 3A). Moreover, colocalization of the early endosomal protein EEA1 and Rab7 was significantly higher in WDR91^Y15N^ cells compared with WDR91^WT^ cells (Figure 3B), indicating persistence of intermediate endosomal compartments, as previously reported in WDR91^KO^ Hela cells^4^. Together, these data show that both p.Gln215* and p.Tyr15Asn variants are associated with delayed endosomal maturation, with the p.Tyr15Asn variant exhibiting a phenotype comparable to the truncating allele under these experimental conditions.

**Figure 3:**
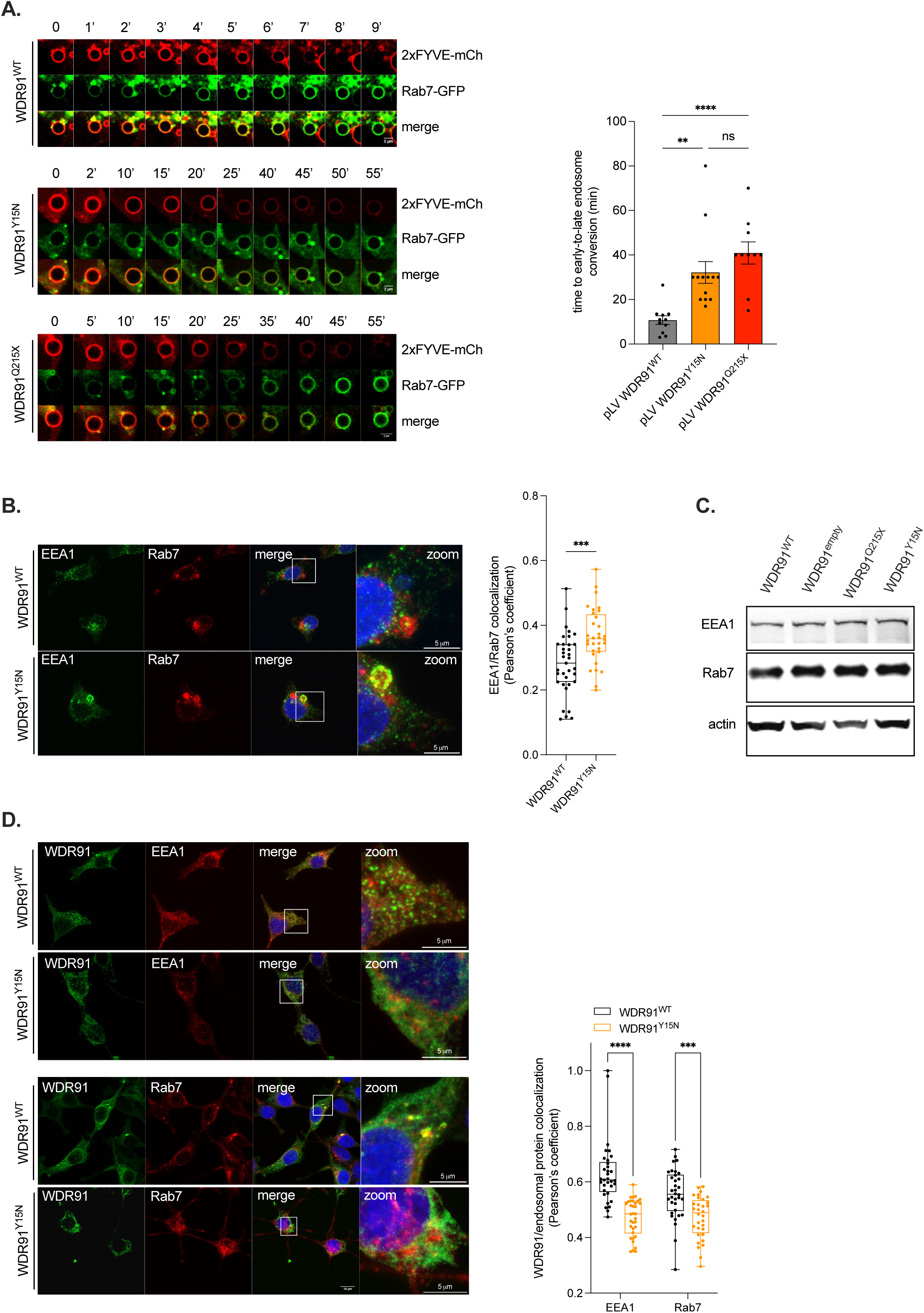
WDR91 p.Gln* and p.Tyr15Asn variants impair early-to-late endosomal maturation. **A.** Time-lapse recording of dynamic changes of the early endosomal marker 2×FYVE-mCherry and the late endosomal protein GFP-Rab7 on endosomes in transduced HEK193 cells (left) and duration of the overlap of 2×FYVE and Rab7 on endosomes (right; one-way ANOVA with Kruskal-Wallis multiple comparison test, ** p<0.01, **** p<10^-4^; at least 10 endosomes per cell line were analyzed). **B.** Colocalization of endogenous EEA1 with Rab7 in WDR91^WT^ and WDR91^Y15N^ cells: representative confocal images (left) and assessment of Pearson’s coefficient (right; Mann-Whitney test, *** p<10^-3^; at least 30 independent cells per cell line were analyzed). **C.** EEA1 and Rab7 protein expression in transduced HEK293 cells, representative blot of three independent experiments. Individual fluorescence channels were analyzed separately due to distinct background signal. **D.** Colocalization of WDR91 with endogenous EEA1 or Rab7 in WDR91^WT^ and WDR91^Y15N^ cells: representative confocal images (left) and assessment of Pearson’s coefficient (right; 2-way ANOVA with Sidak multiple comparison test, *** p<10^-3^, **** p<10^-4^; at least 30 independent cells per cell line were analyzed).

### Colocalization of WDR91 with Rab7-positive endosomes is reduced in WDR91^Y15N^ cells

To further investigate the mechanism underlying delayed endosomal maturation in WDR91^Y15N^ cells, we assessed the subcellular colocalization of WDR91 with the early endosomal marker EEA1 and the late endosomal GTPase Rab7. As previously described, WDR91^WT^ preferentially localized to Rab7-positive vesicles rather than to EEA1-positive early endosomes^4^. In contrast, WDR91^Y15N^ exhibited significantly reduced colocalization with Rab7-positive compartments (Figure 3D). Importantly, EEA1 and Rab7 protein expression levels were comparable across all cell lines (Figure 3C), ruling out altered expression of endosomal markers as a confounding factor.

Altogether, these results indicate that the p.Tyr15Asn variant significantly impairs WDR91 colocalization with Rab7-positive late endosomes, which may contribute to the delayed early-to-late endosomal conversion observed in WDR91^Y15N^ cells.

### WDR91 deficiency promotes accumulation of autophagosomes and autolysosomes in vivo

As WDR91 deficiency leads to accumulation of autophagic cargoes in mouse neurons^5^, we examined whether transcriptional programs related to autophagy were altered in WDR91^KO^ cells compared to CRISPR control cells expressing endogenous WDR91^WT^ protein (hereafter referred to as WDR91^WT/CRISPR^ ^control^ cells) using RNA-seq transcriptomic profiling. Gene set enrichment analysis revealed a significant enrichment of autophagy-related genes in WDR91-deficient cells (NES=1.41, FDR<0.05, q-value=0.03; Figure 4B). Leading-edge genes included multiple factors involved in distinct stages of the autophagy pathway, including initiation and autophagosome nucleation (ATG13, ATG101, AMBRA1 and PIK3R4/VPS15), phagophore elongation (ATG2A, ATG3, ATG4B/C, ATG7, ATG9A/B, ATG12 and ATG16L2), and autophagosome maturation (UVRAG, GABARAP and GABARAPL2), all of which displayed increased expression in WDR91^KO^ cells (Figure 4A). Together, these data indicate transcriptional upregulation of multiple components of the autophagy machinery in WDR91-deficient cells.

**Figure 4:**
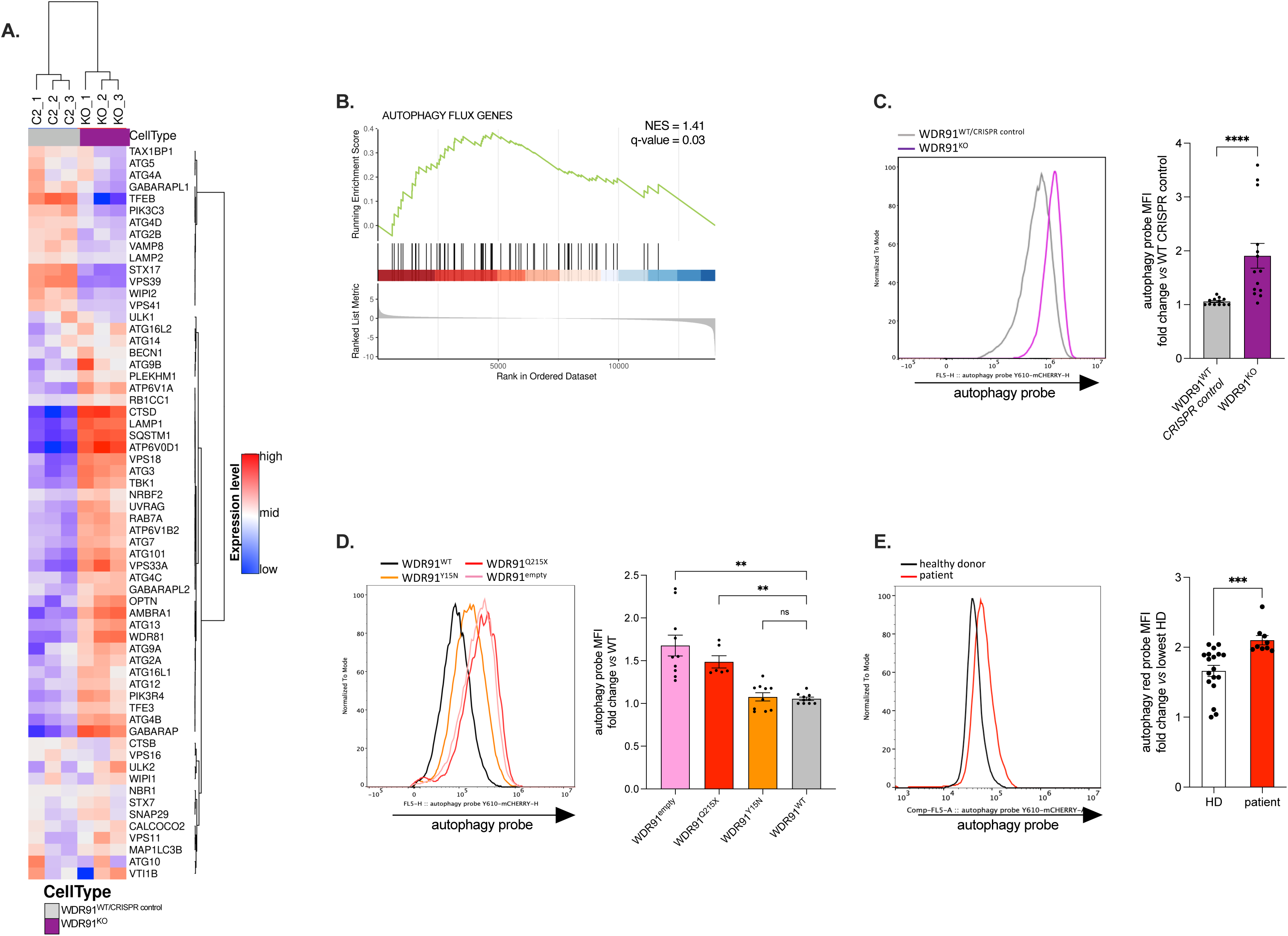
WDR91 deficiency promotes accumulation of autophagosomes and autolysosomes in vivo. **A.** Heat map of significantly differentially-expressed genes encoding autophagic flux proteins in WDR91^WT/CRISPR^ ^control^ and WDR91^KO^ HEK293 cells. Upregulated and downregulated genes are shown in red and blue, respectively. **B.** Gene set enrichment analysis plot of autophagy flux genes and ranked list of the 25 most-contributing genes in WDR91^KO^ cells *versus* WDR91^WT/CRISPR^ ^control^ cells (NES, normalized enrichment score; FDR<0.05). **C-E.** Assessment by flow cytometry of autophagosome accumulation following 2 h-treatment with bafilomycin A_1_ (10 nM) in **C.** WDR91^WT/CRISPR^ ^control^ and WDR91^KO^ HEK293 cells, **D.** WDR91^empty^, WDR91^Q215X^, WDR91^Y15N^ and WDR91^WT^ HEK293 cells and **E.** primary T cells from healthy donors (black line) and patient (red line). Representative flow cytometry histogram of autophagy probe (left). Quantification of geomean of fluorescence intensity (MFI) of autophagy probe (right). (C-E) All data were gathered from four independent experiments including experimental duplicate or triplicate and results were normalized to the lowest control within each experiment. Bars indicate mean ± SEM. Stats: **C** and **E**. Mann Whitney test; **D**. one-way ANOVA with Sidak multiple comparison test. n.s., p>0.05, **p<0.01, *** p<10^-3^, **** p<10^-4^.

To functionally evaluate whether the transcriptional signature observed by RNA-seq translated into alterations of autophagy dynamics, we directly assessed autophagic flux in the HEK293T models by quantifying autophagosome accumulation using a fluorescent probe labeling both autophagosomes and autolysosomes following lysosomal inhibition with bafilomycin A_1_, which blocks autophagosome-lysosome degradation and thereby allows assessment of ongoing autophagic flux.

Consistent with observations in mouse neurons, WDR91^KO^ cells exhibited a significant increase in autophagosomes and autolysosomes compared to WDR91^WT/CRISPR^ ^control^ cells, indicating impaired autophagic turnover (Figure 4C). As expected, expression of the p.Gln215* variant in WDR91^KO^ cells failed to rescue this phenotype, resulting in persistent accumulation of autophagic compartments in WDR91^Q215X^ cells at levels comparable to empty-vector controls, consistent with loss of WDR91 function (Figure 4D). By contrast, expression of the p.Tyr15Asn variant in WDR91^KO^ cells (WDR91^Y15N^ cells) restored autophagic compartment levels to those observed in WDR91^WT^ rescued cells, suggesting that this missense variant does not impair autophagosome and autolysosome turnover under overexpression conditions (Figure 4D). Importantly, WDR91 protein levels remained unchanged after bafilomycin A_1_ treatment, indicating that the comparable autophagic flux observed in WDR91^Y15N^ cells is unlikely to result from stabilization of WDR91 protein secondary to lysosomal degradation blockade (Supplemental Figure 6).

Given the marked reduction of WDR91 protein in primary T lymphoblasts from the patient, we next assessed autophagy in these cells. Patient-derived T cells exhibited significant accumulation of autophagosomes and autolysosomes compared with healthy donor controls, consistent with impaired autophagic turnover *in vivo* (Figure 4E).

Together, these data demonstrate that complete loss of WDR91 disrupts autophagic turnover, whereas the p.Tyr15Asn variant can sustain autophagic compartment dynamics in an overexpression system but is associated with defective autophagy in patient-derived cells.

## Discussion

Here we describe a neurodevelopmental disorder associated with WDR91 deficiency, characterized by severe neurodevelopmental impairment, progressive microcephaly, and structural brain abnormalities including lissencephaly and corpus callosum atrophy. Importantly, our study provides functional evidence linking biallelic *WDR91* variants to defects in endosomal maturation and autophagy, supporting a causal role between WDR91 dysfunction and human disease.

Notably, two independent case reports have described individuals harboring biallelic WDR91 variants and presenting with a highly overlapping neurodevelopmental phenotype, characterized by severe developmental delay, microlissencephaly and agenesis of the corpus callosum^6; 7^. These reports identified two different homozygous splice-site variants of *WDR91*. Although these observations support a recurrent clinical association, both reports remained descriptive and did not include functional validation of the identified variants, and no detailed population frequency data or *in silico* pathogenicity predictions were provided. Our study extends these observations by providing integrated genetic, molecular, and cellular evidence supporting the pathogenicity of *WDR91* variants and delineating the underlying disease mechanism, although genetic evidence remains currently limited to a small number of cases.

Our genetic and functional analyses demonstrate that the p.Gln215* variant results in complete loss of WDR91 protein expression, whereas the p.Tyr15Asn missense variant leads to reduced steady-state protein levels due to increased degradation. Cycloheximide chase experiments indicate accelerated turnover of the WDR91^Y15N^ protein, suggesting reduced stability of the mutant protein. Structural modeling did not predict major global destabilization of the WDR91 fold but instead suggested that the p.Tyr15Asn substitution induces a localized change within the N-terminal region of the protein. Notably, this region encompasses a predicted APC/C-associated D-box degron motif. We therefore hypothesize that the substitution may alter degron accessibility or recognition by the APC/C E3 ubiquitin ligase complex, potentially increasing ubiquitination of Lys44 and thereby facilitating degradation of the mutant protein. However, this mechanism remains to be experimentally validated.

Functionally, we show that both variants impair early-to-late endosomal maturation. Live-cell imaging revealed delayed conversion of PtdIns3P-positive early endosomes to Rab7-positive late endosomes in cells expressing either WDR91^Q215X^ or WDR91^Y15N^. These observations are consistent with the established role of WDR91 in regulating endosomal maturation^4^. WDR91 forms a complex with WDR81 and is recruited to endosomal membranes by the active GTP-bound form of Rab7. Once recruited, the WDR91-WDR81 complex interacts with the Beclin 1-containing class III PI3K complex to modulate Rab7-associated PI3K activity and thereby facilitate early-to-late endosome conversion^4^.

Consistent with this mechanism, we observed reduced colocalization between WDR91^Y15N^ and Rab7. Although our experiments did not directly assess physical interaction between these proteins, the decreased spatial association suggests that the p.Tyr15Asn substitution may impair efficient recruitment of WDR91 to Rab7-positive endosomes. Although the intact C-terminal WD40 repeats of WDR91 are sufficient and necessary to interact with Rab7, the N-terminal region of WDR91 has previously been shown to confer specificity for interaction with the active GTP-bound form of Rab7^4^. Therefore, altered recruitment of the WDR91^Y15N^ protein to Rab7-positive endosomes might contribute to the delayed endosomal maturation phenotype observed in our cellular models.

Our findings are consistent with previous observations in WDR91 conditional knockout mice, in which depletion of Wdr91 impaired early-to-late endosomal conversion in neurons and resulted in defective dendritic arborization and reduced brain size. These mice develop progressive neuronal degeneration after birth, accompanied by accumulation of apoptotic neurons and severe brain atrophy^4^. The clinical presentation of our patient closely parallels this phenotype, with progressive microcephaly and marked corpus callosum hypoplasia. Moreover, the increase in circulating biomarkers of brain injury, including GFAP and neurofilament proteins, suggests ongoing neurodegeneration during early childhood, consistent with the progressive worsening of microcephaly observed in this patient.

WDR91 operates in close functional association with WDR81^4^, another regulator of endosomal trafficking whose deficiency also causes severe neurodevelopmental disease characterized by congenital microcephaly and reduced neocortical gyrification^30^. WDR81 has been shown to regulate endosomal maturation through its interaction with WDR91^3^ and independently participates in autophagic degradation of aggregated proteins through its binding to p62^31^. Experimental studies suggest that the neurodevelopmental phenotype associated with WDR81 deficiency arises primarily from impaired endosomal maturation affecting proliferation of radial glial progenitors during corticogenesis^32^. By analogy, the reduced cortical gyrification observed in our patient may result primarily from the endosomal maturation defects observed in WDR91 mutant cells.

Interestingly, a recent case report described the presence of coarse purplish cytoplasmic granules in neutrophils from a child harboring biallelic WDR81 variants, presenting with severe microcephaly and hypoplasia of the corpus callosum^33^. The recurrence of such cytological abnormalities in disorders involving WDR81 and WDR91 proteins supports a shared pathophysiological framework linking membrane trafficking defects to both neurodevelopmental abnormalities and altered granule homeostasis in hematopoietic cells.

Although extensive analyses of neutrophils were not systematically performed in our study, the presence of abnormal giant granulations in neutrophils in our patient suggests that WDR91 deficiency may also impact vesicular trafficking pathways in myeloid cells, thereby extending the cellular consequences of WDR91 dysfunction beyond the nervous system.

In addition to its role in endosomal trafficking, WDR91 has been shown to regulate autophagy through a distinct molecular mechanism. A previous study reported that WDR91 competes with the HOPS (homotypic fusion and protein sorting) complex component VPS41 for binding to Rab7, thereby modulating late endosome-lysosome fusion events and maintaining appropriate autophagic flux^5^. Through this competition, WDR91 limits excessive Rab7/HOPS association and ensures proper regulation of autophagosome maturation and lysosomal fusion.

Consistent with this dual role of WDR91, our data reveal transcriptional and functional evidence of autophagy dysregulation in WDR91-deficient cells. Transcriptomics analyses showed enrichment of genes involved in multiple stages of the autophagy pathway, including autophagosome initiation, elongation, and maturation. Functional assays further demonstrated impaired autophagic turnover in WDR91^KO^ cells, as indicated by accumulation of autophagosomes and autolysosomes under conditions of lysosomal inhibition. Importantly, a similar accumulation of autophagic compartments was observed in primary T cells derived from the patient, supporting the relevance of this defect *in vivo*.

Interestingly, the p.Tyr15Asn variant was able to restore autophagic compartment dynamics in the overexpression system despite the delayed endosomal maturation observed in these cells. This apparent discrepancy likely reflects supraphysiological expression levels in overexpression systems masking partial loss-of-function effects, as this observation suggests that partial WDR91 activity may be sufficient to sustain Rab7-dependent regulation of autophagic fusion when the mutant protein is expressed at supraphysiological levels. In contrast, patient-derived cells carrying compound heterozygous variants exhibit markedly reduced WDR91 expression and display clear evidence of autophagic disruption, indicating that insufficient WDR91 expression likely contributes to defective autophagic homeostasis *in vivo*. Because the truncating allele abolishes WDR91 expression, patient-derived cells effectively express the p.Tyr15Asn variant alone, providing a physiologically relevant context to assess its functional impact. Taken together, our findings support that p.Tyr15Asn behaves as a partial loss of function allele whose functional impact on autophagy homeostasis depends on protein abundance.

Defects of autophagy have increasingly been recognized as an important pathogenic mechanism in neurodevelopmental disorders. Several monogenic diseases affecting core components or regulators of the autophagy pathway, including Vici syndrome (EPG5 deficiency), ATP6V1A-related disorders, ATG7 deficiency, WIPI2 deficiency, and TECPR2-related disease, share a convergent clinical phenotype characterized by severe developmental delay, microcephaly, brain abnormalities including atrophy/hypoplasia of corpus callosum, and progressive neurodegeneration^34–40^. These disorders arise from defects at different steps of the autophagy pathway, including autophagosome formation (ATG7, WIPI2), lysosomal acidification (ATP6V1A), autophagosome-lysosome fusion (EPG5), or regulation of autophagic trafficking (TECPR2), highlighting the critical requirement for tightly coordinated autophagic flux in neuronal homeostasis. Neurons are indeed particularly dependent on efficient autophagic degradation due to their high metabolic activity and post-mitotic state^41^.

In this context, WDR91 deficiency expands the spectrum of autophagy-related neurodevelopmental disorders by linking Rab7-dependent endosomal maturation to autophagic regulation through a mechanism distinct from canonical autophagy genes. Disruption of autophagic turnover in WDR91 deficiency may therefore contribute to progressive neuronal dysfunction and neurodegeneration, consistent with the worsening microcephaly and increased circulating markers of brain injury observed in our patient. Importantly, the combined impairment of endosomal maturation and autophagy observed in WDR91-deficient cells suggests that defects at the interface between endosomal trafficking and autophagic pathways may represent a distinct pathogenic mechanism within this group of disorders.

Taken together, our findings support WDR91 deficiency as a cause of neurodevelopmental disorder associated with impaired endosomal maturation and altered autophagic homeostasis. These results highlight the importance of coordinated endosomal-autophagic trafficking in brain development and neuronal survival. The presence of abnormal cytoplasmic granulations in neutrophils, also reported in WDR81 deficiency, further suggests that defects in endolysosomal trafficking may extend beyond the nervous system and provide an accessible diagnostic clue.

## Data Availability

All data produced in the present study are available upon reasonable request to the authors

## Acknowledgments

We thank the affected individual and his family for support and motivation. We thank Prof. Geneviève de Saint Basile and Prof. Capucine Picard who provided protocols and inputs for some experiments. We thank Dr. Odile Blanchet who took care of the biological ressources collection and Clément Prouteau who provided administrative support. Ch.M. is supported by a grant from the University Hospital of Angers. This work was supported by French CEREDIH funding.

## Notes

### Competing Interest Statement

The authors have declared no competing interest.

### Clinical Trial

NCT05499091

### Funding Statement

This study was funded by a grant from the University Hospital of Angers and by French CEREDIH funding.

### Author Declarations

Ethics committee of University Hospital of Angers gave ethical approval for this work (ID RCB 2022-A00473-40)

