## Supplementary information for "Biallelic WDR91 variants cause a neurodevelopmental disorder through impaired endosomal maturation and autophagy dysregulation"

**Supplemental Methods**

***Plasmid constructs and cells transfection***

A plasmid encoding Myc-DDK-tagged human WDR91 (pCMV6-Entry, RG205719, Origen) was used as the WT construct. Site-directed mutagenesis was performed using the QuikChange II XL Site-Directed Mutagenesis Kit (Agilent Technologies) to generate the indicated mutant variants. All constructs were sequence-verified prior to use. HEK293T cells were transiently transfected with WDR91 expression plasmids using Lipofectamine 2000 (Thermo Fisher Scientific) according to the manufacturer's instructions. Cells were incubated with transfection complexes for 24 hours before downstream analyses. Stable transfectants were also generated by selection in medium containing G418 (neomycin 1 mg/mL, Sigma-Aldrich) following transfection. Resistant clones were expanded under continuous antibiotic selection and screened for WDR91 expression by immunoblotting and immunofluorescence. We used 1 µg of plasmid DNA to transfect $5.10^5$ cells unless otherwise stated in figure legends.

***RT-qPCR analysis of mRNA expression***

Total RNA was isolated using RNeasy Mini kit (Qiagen), treated with DNase (RNase-Free DNase Set, Qiagen), and reverse transcribed to generate cDNA with High-Capacity RNA-to-cDNA™ Kit (Themo-Fisher Scientific) according to manufacturer's instructions. The cDNA was subjected to qPCR using Power SYBR Green kit (Applied Biosystems) and primers specific for *beta-actin*, *RPS18*, *EF1a* and *WDR91* (Integrated DNA Technologies, supplementary Table 1). Relative expression was calculated using the ddCt method, using *actin, RPS18* and *EF1a* as a housekeeping genes, and relevant calibrator sample (explained in figure legends for respective samples).

***Bioinformatics analyses***

*Quantification of gene.* Expression STAR was used to obtain the number of reads associated to each gene in the Gencode v47 annotation (restricted to protein-coding genes, antisense and lincRNAs)[1]. Raw counts for each sample were imported into R statistical software. Extracted count matrix was normalized for library size and coding length of genes to compute FPKM expression levels.

*Unsupervised analysis.* The Bioconductor edgeR package was used to import raw counts into R statistical software, and compute normalized log2 CPM (counts per millions of mapped reads) using the TMM (weighted trimmed mean of M-values) as normalization procedure. The normalized expression matrix from a custom gene list was used to classify the samples according to their gene expression patterns using hierarchical clustering. Hierarchical clustering was performed by stats::hclust function (with euclidean distance and ward.D method).

*Differential expression analysis*. The Bioconductor edgeR package was used to import raw counts into R statistical software. Differential expression analysis was performed using the Bioconductor limma package and the voom transformation. To improve the statistical power of the analysis, only genes expressed in at least one sample (FPKM >= 0.1) were considered. A qval threshold of <= 0.01 and a minimum fold change of 2 were used to define differentially expressed genes.

*Pathway enrichment analysis – GSEA*. Gene list from the differential analysis was ordered by decreasing log2 fold change. Gene set enrichment analysis was performed by clusterProfiler::GSEA function using the fgsea algorithm. Gene sets from MSigDB v666 database were selected among the H classes, keeping only gene sets defined by 666 genes.

**Supplementary Table 1:** qPCR primer sequences

| **Gene** | **Forward** | **Reverse** |
|---|---|---|
| *WDR91* | 5'-GGAGCGCACTGACGAGC-3' | 5'-TCTTATCCACCCGGAACCCC-3' |
| *Beta-actin* | 5'-CCAGATCATGTTTGAGACCT-3' | 5'-GTTCCACCTCATCCTCAGTG-3' |
| *RPS18* | 5'-CCTTTGCCATCACTGCCA-3' | 5'-GGCATACCCCTCGTAGAT-3' |
| *EF1a* | 5'-TGATATGGTTCCTGGCAAGC-3' | 5'-TAGCCTTCTGAGCTTTCTGG-3' |

### Supplemental Figure S1

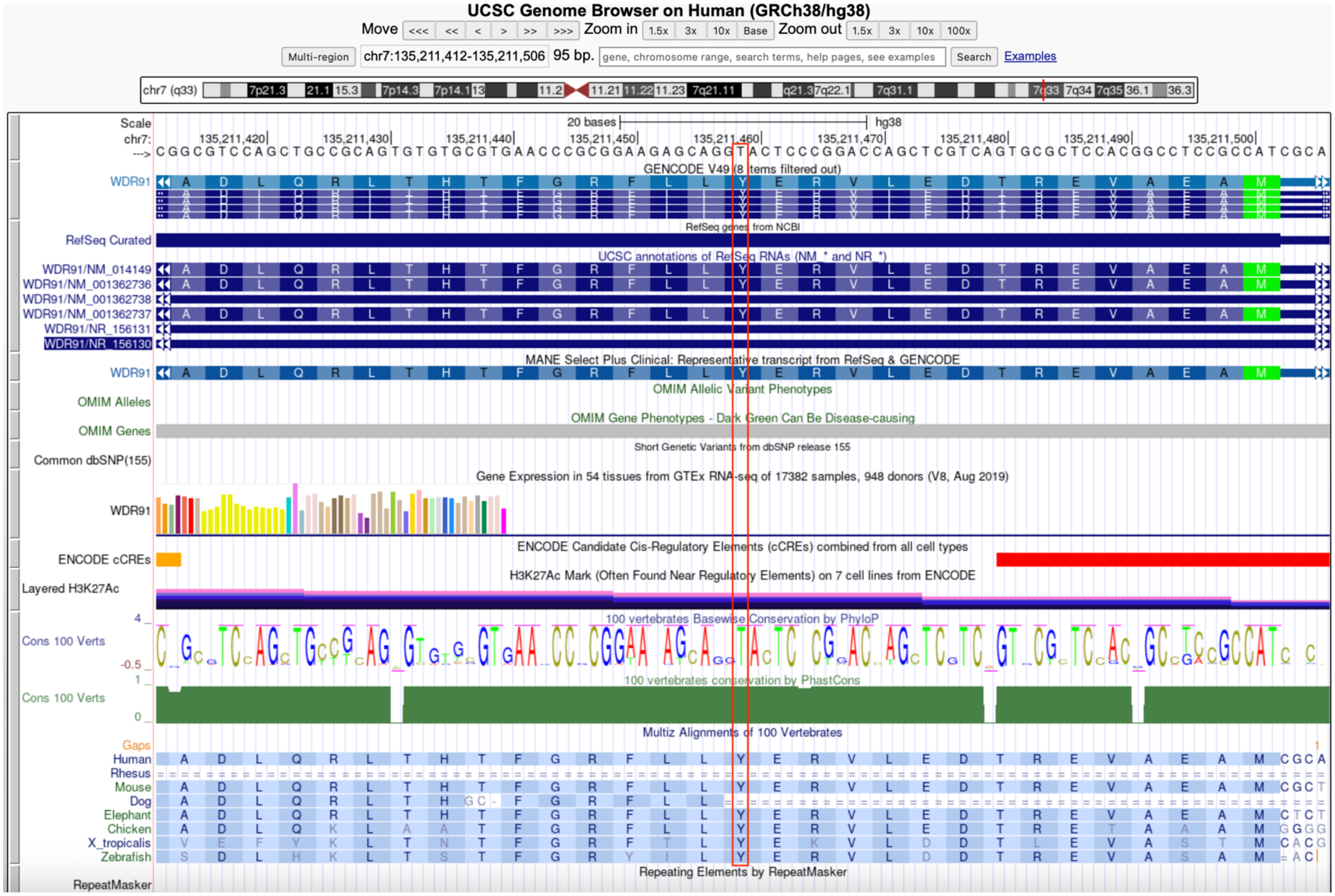

**Supplemental Figure S1:** Evolutionary conservation at the nucleotide level was assessed using PhyloP and PhastCons scores from the UCSC Genome Browser (100-way vertebrate alignment). c.43T is highlighted in red.

**Supplemental Figure S2:** Tri-dimensional AlphaFold-predicted WDR91 protein structure. **A.** WDR91$^{WT}$: file WDR91_WT_rotation.mp4; **B.** WDR91$^{Y15N}$: file WDR91_Y15N_rotation.mp4. Amino acid 15 residue is highlighted in red, known and predicted domains are highlighted in different colors as follows: N-terminal domain (1-55) in pink, predicted degron in dark blue, predicted ubiquitinylated Lysine 44 in orange, coil-coiled domain in cyan, SAM domain in yellow and WD40 domains in green. See corresponding supplementary files

**Supplemental Figure S3**

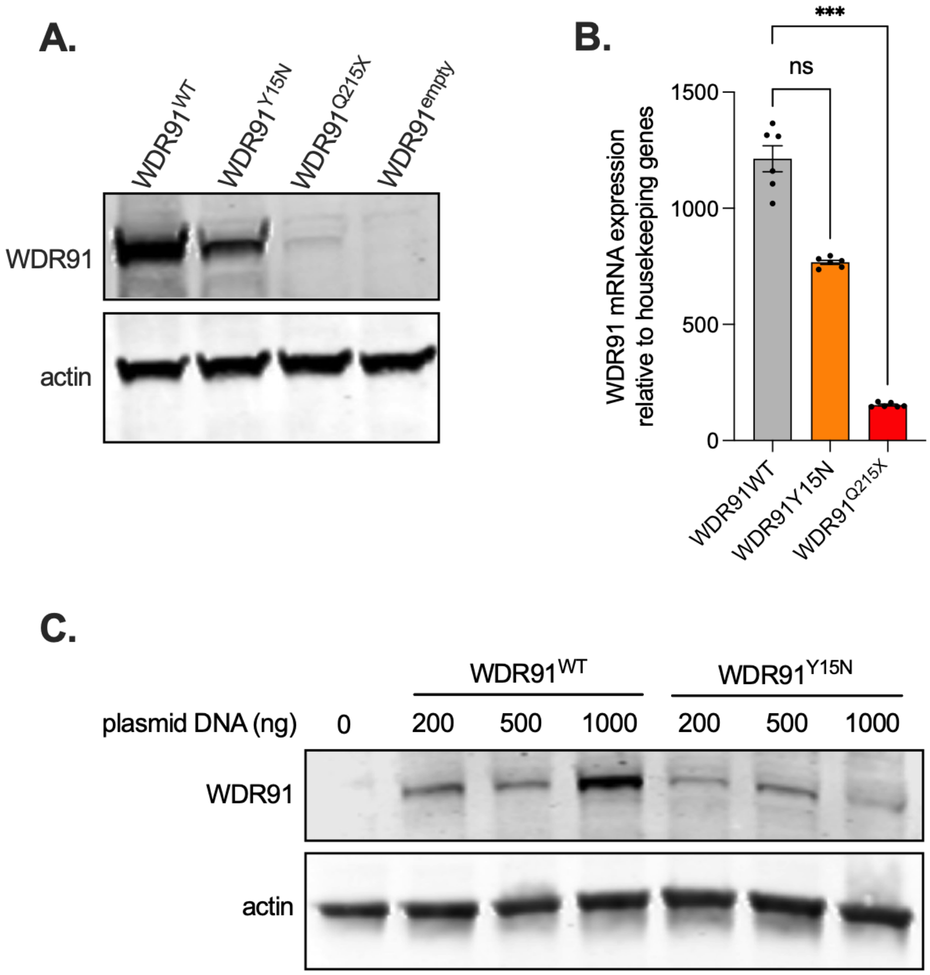

**Supplemental Figure S3: A.** WDR91 protein expression in HEK293 $WDR91^{KO}$ cells transfected with an empty plasmid ($WDR91^{empty}$) or a plasmid encoding WT ($WDR91^{WT}$), p.Gln215* ($WDR91^{Q215X}$), or p.Tyr15Asn ($WDR91^{Y15N}$) WDR91 cDNA. Representative blot within 3 independents experiments. **B.** Quantification WDR91 mRNA expression relative to housekeeping genes in HEK293 WDR91KO cells transduced with an empty lentiviral vector ($WDR91^{empty}$) or a vector encoding WT ($WDR91^{WT}$), p.Gln215* ($WDR91^{Q215X}$), or p.Tyr15Asn ($WDR91^{Y15N}$) WDR91 cDNA. All data were gathered from three independent experiments including experimental duplicate. Bars indicate mean +/- SEM. Stats: one-way ANOVA with Dunn's multiple comparison test. n.s. $p>0.05$, *** $p<10^{-3}$. **C.** WDR91 protein expression in CHO cells transfected with increasing doses of a plasmid encoding WT ($WDR91^{WT}$) or p.Tyr15Asn ($WDR91^{Y15N}$) WDR91 cDNA. Representative blot from 3 independents experiments. For Western blots, individual fluorescence channels were analyzed separately due to distinct background signal.

**Supplemental Figure S4**

**A.**

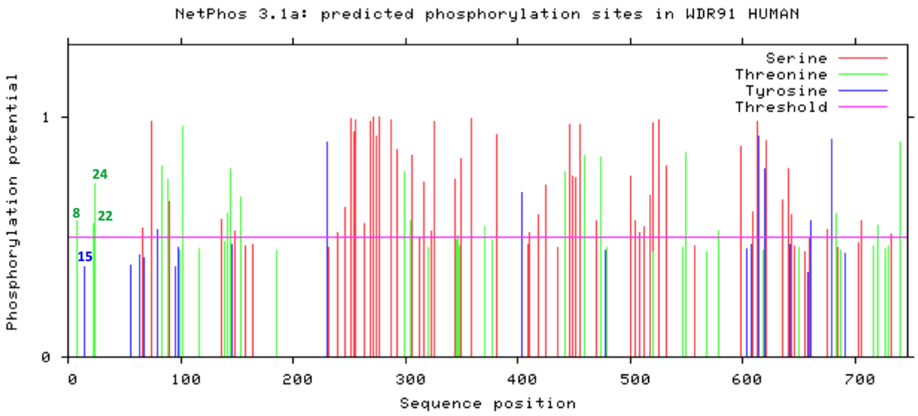


**B.**

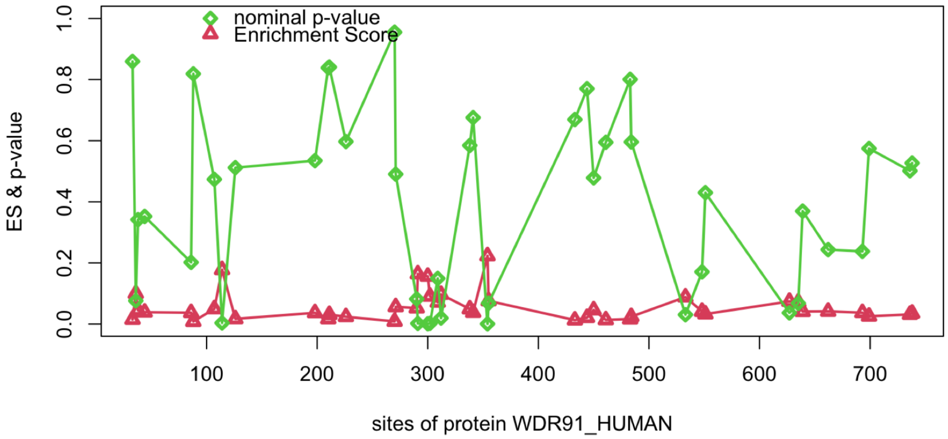


**C.**

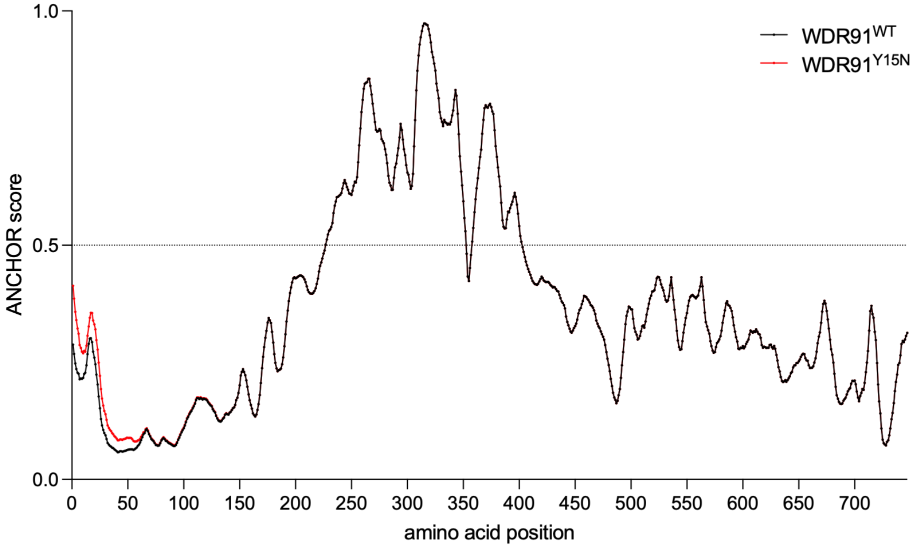

**Supplemental Figure S4: A.** Putative phosphorylation sites on the full-length WDR91 protein sequence (NetPhos 3.1, default parameters). A NetPhos score > 0.5 was considered as significant (pink line). **B.** Putative lysine acetylation candidates on the full-length WDR91 protein sequence under acetyltransferase GCN5/PCAF model (ASEB, default parameters). **C.** Quantification of intrinsic disorder and disorder-mediated binding propensity of $WDR91^{WT}$ (black line) and $WDR91^{Y15N}$ (red line) through ANCHOR score using IUPred2A (long mode) and ANCHOR2 under identical parameters for WT and p.Tyr15Asn WDR91 sequences.

**Supplemental Figure S5**

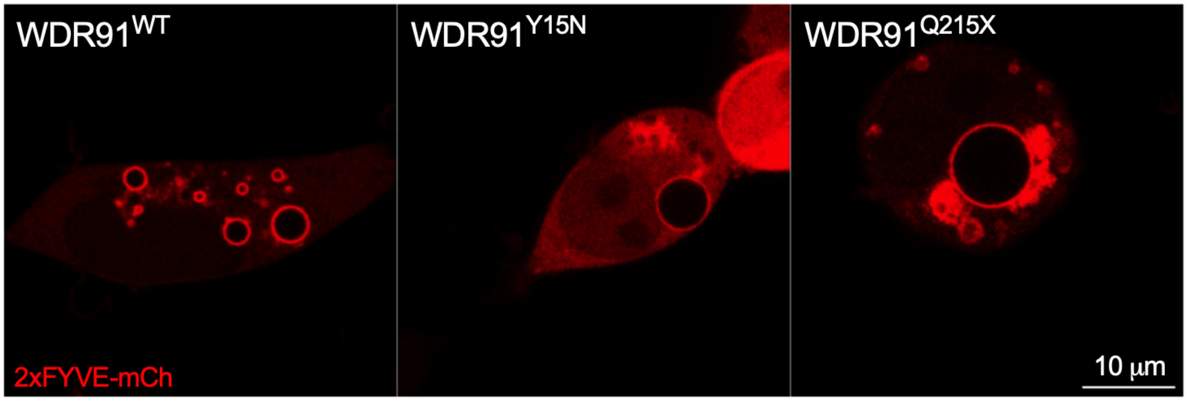

**Supplemental Figure S5:** Immunofluorescence staining of $WDR91^{WT}$ (left), $WDR91^{Y15N}$ (middle) and $WDR91^{Q215X}$ (right) HEK 293T cells with the 2xFYVE-mCherry probe revealing enlarged early endosomes in $WDR91^{Y15N}$ and $WDR91^{Q215X}$ cells compared to $WDR91^{WT}$ cells. 2xFYVE probe specifically binds to phosphatidylinositol 3-phosphate (PtdIns3P) and is a marker of early endosomes[2]. Representative fields within more than 30 independent acquisitions for each cell lines.

**Supplemental Figure S6**

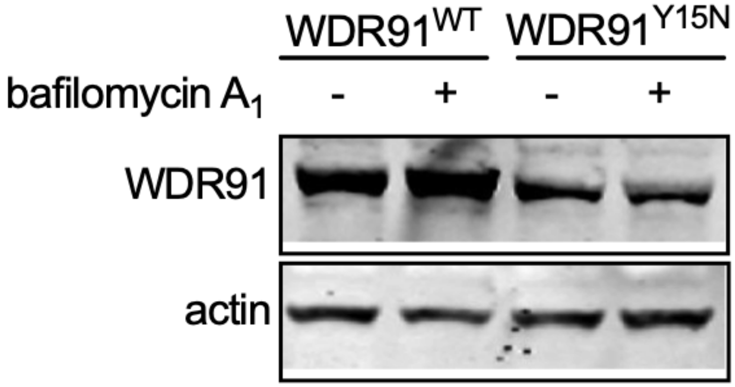

**Supplemental Figure S6:** WDR91 protein expression in HEK 293T $WDR91^{WT}$ and $WDR91^{Y15N}$ cells following 2 hours-treatment with bafilomycin $A_1$ 10 nM. Representative blot within 3 independents experiments. For Western blots, individual fluorescence channels were analyzed separately due to distinct background signal.
